# Age-dependent heterogeneity of lymph node metastases and survival identified by analysis of a national breast cancer registry

**DOI:** 10.1101/2021.01.21.21250252

**Authors:** Michael Behring, Prachi Bajpai, Farrukh Afaq, Amr Elkholy, Hyung-Gyoon Kim, Sameer Al Diffalha, Sadeep Shrestha, Upender Manne

## Abstract

**Background:** For several cancers, including those of the breast, young age at diagnosis is associated with an adverse prognosis. Although this effect is often attributed to heritable mutations such as BRCA1/2, the relationship between pathologic features, young age of onset, and prognosis for breast cancer remains unclear. In the present study, we highlight links between age of onset and lymph node metastasis (NM) in US women with breast cancer.

**Methods:** Case listings from Surveillance, Epidemiology, and End Result (SEER) 18 registry data for women with breast cancer, which include information on race, were used. NM and its associated outcomes were evaluated for a subset of women with receptor subtype information and then compared against a larger, pre-subtype validation set of data from the same registry. Age of diagnosis was a 5-category variable; under 40 years, 40-49 years, 50-59 years, 60-69 years and 70+ years. Univariate and adjusted multivariate survival models were applied to both sets of data.

**Results:** As determined with adjusted logistic regression models, women under 40 years old at diagnosis had 1.55 times the odds of NM as women 60-69 years of age. The odds of NM for (HR = hormone receptor) HR+/HER2+, HR-/HER2+, and triple-negative breast cancer subtypes were significantly lower than those for HR+/HER2-. In subtype-stratified adjusted models, age of diagnosis had a consistent trend of decreasing odds of NM by age category, most noticeable for HR+ subtypes of luminal A and B. Univariate 5-year survival by age was worst for women under 40 years, with NM attributable for 49% of the hazard of death from cancer in adjusted multivariate models.

**Conclusions:** Lymph node metastasis is age-dependent, yet not all molecular subtypes are clearly affected by this relationship. For <40-yr-old women, NM is a major cause for shorter survival. When stratified by subtype, the strongest associations were in HR+ groups, suggesting a possible hormonal connection between young age of breast cancer onset and NM.

## Background

In 2020, an estimated 276,480 new cases of breast cancer were diagnosed in the United States (https://seer.cancer.gov/statfacts/html/breast.html). For women with breast cancer, the infiltration of tumor cells into surrounding lymph nodes is associated with a poor prognosis. Lymph node metastasis (NM), a means for the regional and distant spread of tumor cells, has a considerable influence upon treatment options and patient survival. Approximately 60% of all newly diagnosed cases of breast cancer are localized (non-metastatic). However, one third of the patients with localized cancers will eventually develop metastatic disease [1]. Of all new cases of breast cancer, another third have regional NM at the time of diagnosis [1]. Lymph nodes usually represent the first site of metastasis of breast cancer, and they initiate the process of metastasis of the disease.

Breast cancer is distinctive among highly prevalent cancers in that women with a young age of onset often have a more aggressive form of the disease. Although younger women are eligible for more intensive therapy, they nevertheless have, relative to older patients, worse survival and a higher recurrence rate [2,3]. Although links between molecular/receptor subtypes and disease progression (metastasis in particular) are commonplace [4–6], there has been little research into the durability of these relationships across age groups. Estrogen receptor-positive status has an unclear prognostic influence across ages [7–9]. However, *ERBB2*/*HER2* receptor status seems to be more frequent and is associated with a lower survival of younger women [10]. A few negatively associated prognostic variables for early onset breast cancer are identified [11–13], yet the connection between patient characteristics and metastasis has not been fully assessed. In the present study, we used nation-wide data from Surveillance Epidemiology and End Result (SEER) cancer registries to gain a better understanding of the relationship between lymph node metastasis, receptor subtype, and age-dependent patient characteristics of breast cancer.

## Materials and Methods

### Study population data

The present analysis involved data from the SEER 18 SEER*Stat8.3.8 database case listings. The data in SEER 18 represents 27.8% of the total US population and 18 cancer registries [14]. After excluding patients with non-ductal or lobular tumor histology, male gender, missing information on lymph node metastasis, and follow up of less than 6 months, our final sample consisted of 717,331 women diagnosed with breast cancer from 1975-2017. Since SEER did not start recording information regarding the HER2 receptor subtype until 2010, we used a subset of the overall data that offered complete receptor subtype analysis. The receptor subtype set of data is made up of 223,986 cases of breast cancer with first and only primary tumor for patients diagnosed for the ∼ 7 year period of 2010-2017.

### Study design

The present study used both case-control and follow-up designs. Cases were defined as breast cancer patients with NM at the time of diagnosis. Controls were patients with no NM at diagnosis. Data for all eligible patients were used, and no matching of controls to cases was done. A response variable of lymph node metastasis was defined as a binary outcome using AJCC 7^th^ edition tumor, nodes, and metastases (TNM) staging [15]. This large data set with NM information is consistent with AJCC 6^th^ edition TNM staging [16]. All nodes (N) values of N0 (including N0 (i-) and N0 (mol-)) were considered NM negative, and any other N values (N1-N4) were considered positive. Included were patient demographic measures of age and ethnicity as well as tumor differentiation and staging information. Breast cancer subtype was based on receptor status. The “HR” abbreviation for hormone receptor represents both estrogen (ER) and progesterone (PR) receptor status. HER2 indicates human epidermal growth factor 2 receptor status. Borderline information on HER2 status was excluded from analysis of SEER*Stat data queries [17]. Age of onset was considered as both a continuous and a categorical variable. Categories of age were based on preceding literature [8,18]: under 40 years, 40-49 years, 50-59 years, 60-69 years, and 70+ years. Additional survival analysis was accomplished with time to death from cancer as an outcome (Tables 1a and 1b).

**Table 1a.** Clinical and demographic characteristics of US women with breast cancer by lymph node status (SEER, 2010-2017)

| **Lymph node metastasis**<br>**Characteristics** | **negative**<br>**(N=151827)** | **positive**<br>**(N=72159)** |
| --- | --- | --- |
| Breast tumor by receptor subtype |  |  |
| - HR-/HER2- (triple-negative) | 18418 (12.1%) | 9189 (12.7%) |
| - HR-/HER2+ | 6819 (4.5%) | 4927 (6.8%) |
| - HR+/HER2- | 110069 (72.5%) | 47318 (65.6%) |
| - HR+/HER2+ | 16521 (10.9%) | 10725 (14.9%) |
| Age (years) | 60.5 ± 12.9 | 56.9 ± 13.3 |
| Age categories |  |  |
| - <40 | 7342 (4.8%) | 6702 (9.3%) |
| - 40-49 | 25117 (16.5%) | 15678 (21.7%) |
| - 50-59 | 38185 (25.2%) | 19930 (27.6%) |
| - 60-69 | 42631 (28.1%) | 16746 (23.2%) |
| - 70+ | 38552 (25.4%) | 13103 (18.2%) |
| Cause of death |  |  |
| - alive | 142734 (94.0%) | 60886 (84.4%) |
| - breast | 4014 (2.6%) | 8767 (12.1%) |
| - other | 5079 (3.3%) | 2506 (3.5%) |
| Race |  |  |
| - White | 120218 (79.2%) | 54391 (75.4%) |
| - American Indian/Alaska Native | 925 (0.6%) | 512 (0.7%) |
| - Asian or Pacific Islander | 14083 (9.3%) | 6811 (9.4%) |
| - African American | 15499 (10.2%) | 10011 (13.9%) |
| - Unknown | 1102 (0.7%) | 434 (0.6%) |
| Metastasis (AJCC M 7<sup>th</sup> ed. 2010) |  |  |
| - No distant metastasis | 138047 (98.8%) | 60336 (90.3%) |
| - Distant metastasis | 1737 (1.2%) | 6485 (9.7%) |
| Tumor size (AJCC T 7<sup>th</sup> ed.2010) |  |  |
| - T1 (<2 cm) | 98954 (71.7%) | 22125 (33.8%) |
| - T2 (2 cm-5 cm) | 33586 (24.3%) | 29815 (45.6%) |
| - T3 (>5 cm) | 3788 (2.7%) | 7631 (11.7%) |
| - T4 (extension into chest wall/skin) | 1727 (1.3%) | 5798 (8.9%) |
| Grade |  |  |
| - Well differentiated; Grade I | 38066 (25.1%) | 7535 (10.4%) |
| - Moderately differentiated; Grade II | 63272 (41.7%) | 29342 (40.7%) |
| - Poorly differentiated; Grade III | 45972 (30.3%) | 32305 (44.8%) |
| - Undifferentiated; anaplastic; Grade IV | 340 (0.2%) | 244 (0.3%) |
| - Unknown | 4177 (2.8%) | 2733 (3.8%) |

**Table 1b.** Clinical and demographic characteristics of US women with breast cancer by lymph node status (SEER 1975-2017)

| **Lymph node metastasis**<br>**Characteristics** | **negative**<br>**(N=459656)** | **positive**<br>**(N=257675)** |
| --- | --- | --- |
| Age (years) | 60.7 ± 13.5 | 57.3 ± 13.8 |
| Age categories |  |  |
| - <40 | 24485 (5.3%) | 24526 (9.5%) |
| - 40-49 | 80917 (17.6%) | 57697 (22.4%) |
| - 50-59 | 110958 (24.1%) | 67060 (26.0%) |
| - 60-69 | 112844 (24.5%) | 54294 (21.1%) |
| - 70+ | 130452 (28.4%) | 54098 (21.0%) |
| Cause of death |  |  |
| - alive | 332223 (72.3%) | 141349 (54.9%) |
| - breast | 36964 (8.0%) | 77168 (29.9%) |
| - other | 90469 (19.7%) | 39158 (15.2%) |
| Race |  |  |
| - White | 378616 (82.4%) | 204948 (79.5%) |
| - American Indian/Alaska Native | 2316 (0.5%) | 1514 (0.6%) |
| - Asian or Pacific Islander | 35863 (7.8%) | 19267 (7.5%) |
| - African American | 40497 (8.8%) | 30963 (12.0%) |
| - Unknown | 2364 (0.5%) | 983 (0.4%) |
| Grade |  |  |
| - Well differentiated; Grade I | 94056 (20.5%) | 20760 (8.1%) |
| - Moderately differentiated; Grade II | 174711 (38.0%) | 87336 (33.9%) |
| - Poorly differentiated; Grade III | 131972 (28.7%) | 105851 (41.1%) |
| - Undifferentiated; anaplastic; Grade IV | 4847 (1.1%) | 4105 (1.6%) |
| - Unknown | 54070 (11.8%) | 39623 (15.4%) |

### Statistical analyses

In depicting the univariate relationship between NM, receptor subtype, and individual covariates, we applied chi-square tests for categorical, and t-tests for continuous p-values. Logistic regression models were constructed for each variable to obtain univariate odds of NM and 95% confidence intervals. To examine whether the effect of age upon odds of NM was modified by receptor subtype, we performed 4 separate, adjusted logistic regression analyses stratified by subtype. Logistic regression modeling was also used to adjust for potential confounding variables, both for full data set and in receptor-based subtype subset analyses. Kaplan-Meier log-rank tests and Cox proportional hazard models were used to estimate the effect of age upon survival and NM. We measured 5-year survival for the similar age categories used in the analyses of Alteri et al. [19]. For 5 separate age category-stratified survival models, we calculated the proportion of hazard of death that was attributable to NM (attributable fraction analysis) by comparing a counterfactual survival function (excluding NM from baseline) to a factual survival function, including NM [20,21]. In Cox models, variables adjusted for were NM, race, size of tumor, and receptor subtype. All statistical analyses utilized R version 3.6.2 (2019-12-12) [22].

## Results

### Overall study population

Associations with NM were consistent across overall (1975-2017) and subset (2010—2017) analyses and showed that, at diagnosis, 32-36% of women had nodal metastasis of breast cancer. In follow up, women with NM made up a larger proportion of breast cancer deaths, were more African American, had tumors of larger size at diagnosis, and had more distant metastases at diagnosis (Table 1a and 1b). Additional information unique to the test data set also showed higher grade/more poorly differentiated tumors for NM cases. Women with nodal metastases were also younger than controls (non-NM), with an average age at diagnosis of 57 years (61 years for controls). Lastly, a larger proportion of women had HR+/HER2+, triple-negative, and HR-/HER2+ subtype cancers than controls. See Tables 1a and 1b for a description of patient variables across nodal metastasis outcomes. In a subset analysis (data not shown) of women under 40 years in 1975-2017 data, no effect modification by age was found for the association between race, grade, and NM.

### Odds of nodal metastasis

The adjusted odds of NM for common variables in both datasets were available for race, age, tumor size, and tumor grade. The relationship between age and the adjusted odds of NM had similar estimates for both sets of data. Among all age groups, women under 40 had the highest adjusted odds of NM (1.55 in subset and 1.74 in overall data). Both sets of data confirmed tumor size as being strongly associated with NM. Relative to Whites, African American women had higher adjusted odds of NM (1.13 in subset and 1.23 in overall data). Higher tumor grades were also positively associated with NM, with the effect increasing parallel to the loss of differentiation. See Tables 2a and 2b for details of univariate and adjusted odds of NM.

**Table 2a.** Odds of nodal metastasis, unadjusted and adjusted models (SEER, 2010-2017)

| Odds of lymph node metastasis<br>Characteristics | Unadjusted |  | Adjusted |  |
| --- | --- | --- | --- | --- |
|  | OR | 95% CI | OR | 95% CI |
| Breast tumor by receptor subtype |  |  |  |  |
| - HR-/HER2- (triple negative) | 1.16 | 1.13-1.19 | 0.60 | 0.58-0.62 |
| - HR-/HER2+ | 1.68 | 1.62-1.75 | 0.87 | 0.83-0.91 |
| - HR+/HER2- | 1 | ref | 1 | ref |
| - HR+/HER2+ | 1.51 | 1.47-1.55 | 0.94 | 0.91-0.97 |
| Age categories |  |  |  |  |
| - <40 | 2.32 | 2.24-2.41 | 1.55 | 1.49-1.62 |
| - 40-49 | 1.59 | 1.55-1.63 | 1.37 | 1.33-1.41 |
| - 50-59 | 1.33 | 1.30-1.36 | 1.21 | 1.18-1.25 |
| - 60-69 | 1 | ref | 1 | ref |
| - 70+ | 0.87 | 0.84-0.89 | 0.80 | 0.77-0.82 |
| Race |  |  |  |  |
| - White | 1 | ref | 1 | ref |
| - American Indian/Alaska Native | 1.22 | 1.10-1.36 | 1.08 | 0.95-1.23 |
| - Asian or Pacific Islander | 1.07 | 1.04-1.10 | 0.92 | 0.88-0.95 |
| - African American | 1.43 | 1.39-1.47 | 1.13 | 1.09-1.17 |
| - Unknown | 0.85 | 0.74-0.98 | 0.87 | 0.76-0.99 |
| Metastasis (AJCC M 7<sup>th</sup> ed. 2010) |  |  |  |  |
| - No distant metastasis | 1 | ref | 1 | ref |
| - Distant metastasis | 8.54 | 8.10-9.02 | 3.68 | 3.46-3.91 |
| Tumor size (AJCC T 7<sup>th</sup> ed. 2010) |  |  |  |  |
| - T1 (<2 cm) | 1 | ref | 1 | ref |
| - T2 (2 cm-5 cm) | 3.97 | 3.89-4.06 | 3.40 | 3.32-3.48 |
| - T3 (>5 cm) | 9.01 | 8.64-9.39 | 7.00 | 6.71-7.32 |
| - T4 (extension into chest wall/skin) | 15.02 | 14.21-15.88 | 9.91 | 9.34-10.51 |
| Grade |  |  |  |  |
| - Well differentiated; Grade I | 1 | ref | 1 | ref |
| - Moderately differentiated; Grade II | 2.34 | 2.28-2.41 | 1.70 | 1.65-1.76 |
| - Poorly differentiated; Grade III | 3.55 | 3.45-3.65 | 2.00 | 1.94-2.08 |
| - Undifferentiated; anaplastic; Grade IV | 3.63 | 3.07-4.29 | 1.97 | 1.62-2.38 |
| - Unknown | 3.31 | 3.13-3.49 | 1.62 | 1.52-1.74 |

**Table 2b.** Odds of nodal metastasis, unadjusted and adjusted models (SEER 1975-2017)

| **Odds of lymph node metastasis**<br>**Characteristics** | **Unadjusted** |  | **Adjusted** |  |
| --- | --- | --- | --- | --- |
|  | **OR** | **95% CI** | **OR** | **95% CI** |
| Age categories |  |  |  |  |
| - <40 | 2.08 | 2.04-2.12 | 1.74 | 1.71-1.78 |
| - 40-49 | 1.48 | 1.46-1.50 | 1.39 | 1.37-1.41 |
| - 50-59 | 1.26 | 1.24-1.27 | 1.22 | 1.20-1.23 |
| - 60-69 | 1 | ref | 1.00 | ref |
| - 70+ | 0.86 | 0.85-0.87 | 0.87 | 0.85-0.89 |
| Race |  |  |  |  |
| - White | 1 | ref | 1 | ref |
| - American Indian/Alaska Native | 1.21 | 1.13-1.28 | 1.14 | 1.06-1.21 |
| - Asian or Pacific Islander | 0.99 | 0.97-1.01 | 0.92 | 0.91-0.94 |
| - African American | 1.41 | 1.39-1.43 | 1.23 | 1.21-1.25 |
| - Unknown | 0.77 | 0.71-0.83 | 0.76 | 0.70-0.81 |
| Grade |  |  |  |  |
| - Well differentiated; Grade I | 1 | ref | 1 | ref |
| - Moderately differentiated; Grade II | 2.26 | 2.23-2.30 | 2.20 | 2.17-2.24 |
| - Poorly differentiated; Grade III | 3.63 | 3.57-3.70 | 3.31 | 3.18-3.57 |
| - Undifferentiated; anaplastic; Grade IV | 3.84 | 3.67-4.01 | 3.53 | 3.38-3.69 |
| - Unknown | 3.32 | 3.25-3.39 | 3.23 | 3.17-3.30 |

### Receptor subtype

In receptor-stratified data, adjusted estimates of NM by subtype showed that TNBC cancers had lower odds of nodal metastasis relative to the HER2-/HR+ receptor subtype (OR 0.74, 95% CI 0.70-0.78) (Table 2b). There was, however, no significant increase in the odds of having NM for either HER2+ subtypes of HER2+/ER+ or HER2+/ER- (Table 2a). There was a young-to-old gradient in the odds of NM, which is most apparent in for HR+ subtypes of tumors, HER2-/ER+ and lHER2+/ER+ (Figure 1). In HR-/HER2+subtypes, age at diagnosis had a similar trend in odds estimates per age group, but showed no significant difference between ages <40-59. In addition, triple-negative subtypes showed a slightly higher association with NM for those younger than 60 but with no age gradient (Figure 1).

**Figure 1.**
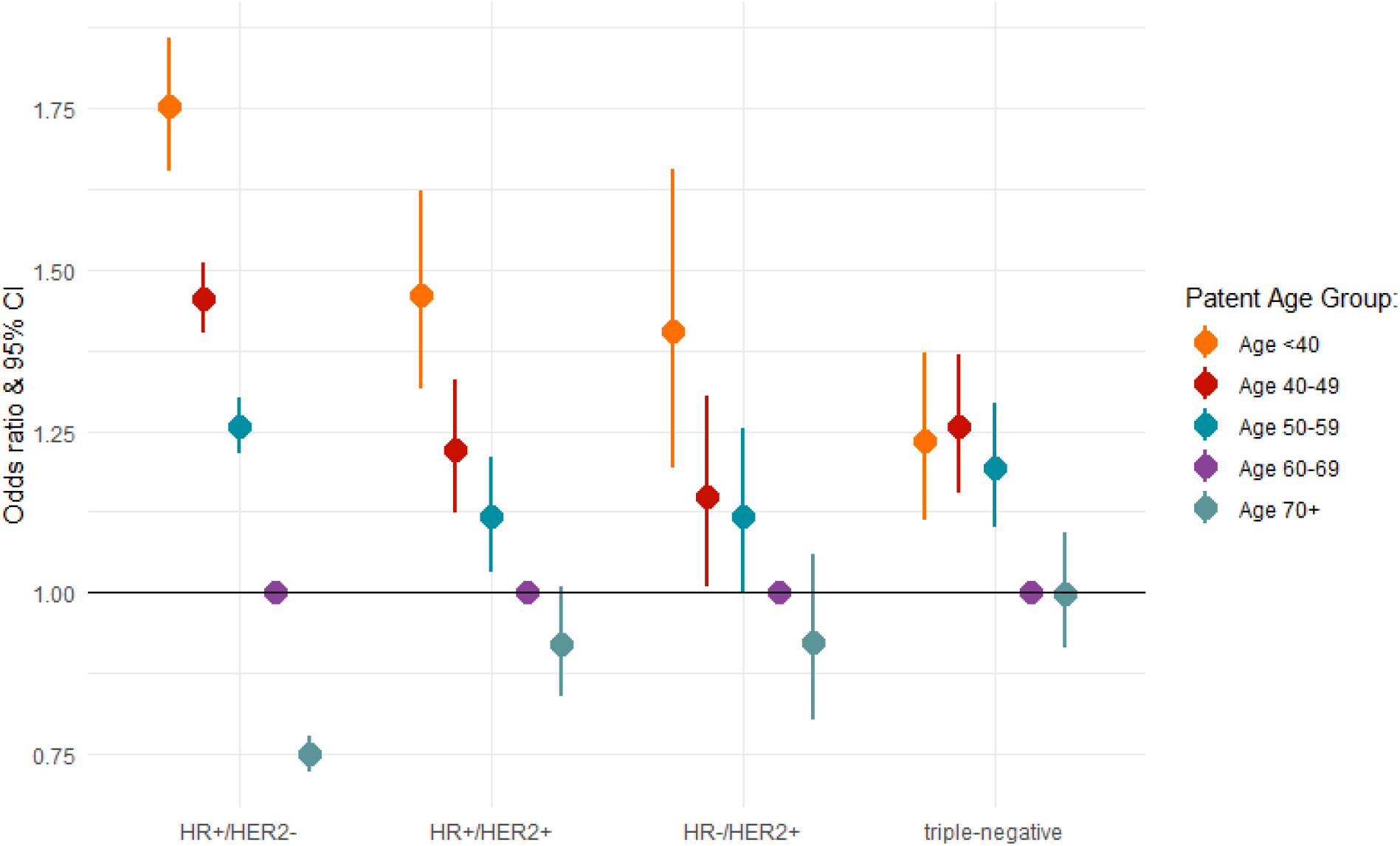
Comparison of adjusted (M stage, T stage, grade) odds of nodal metastasis by age, stratified by receptor subtype, SEER 2010-2017 (ref. group age 60-69 years). Adjusted for race and tumor size.

#### Survival analysis

For both subset and overall data, Kaplan-Meier analyses consistently showed women under 40 as sharing the worst survival outcomes with women aged 70 years and above. Although women 70 years and older had low survival, women under 40 years old began follow-up with survival similarly high with other age categories (40-69 years), but there was a decline starting at ∼15 months which surpassed 70-year-olds at ∼22 months. Women under 40 years of age had the lowest probability of 5-year survival at 0.87, with age groups from 40-69 having similar 5-year survivals at approximately 0.91-0.93, with the value for the oldest group of women slightly decreasing to 0.89. As determined with adjusted Cox analyses, in both sets of data, women younger than 40 also had a higher fraction of hazard of death from cancer (∼50% vs. ∼37% at start) due to NM than women 70 years and above (Figure 2).

**Figure 2.**
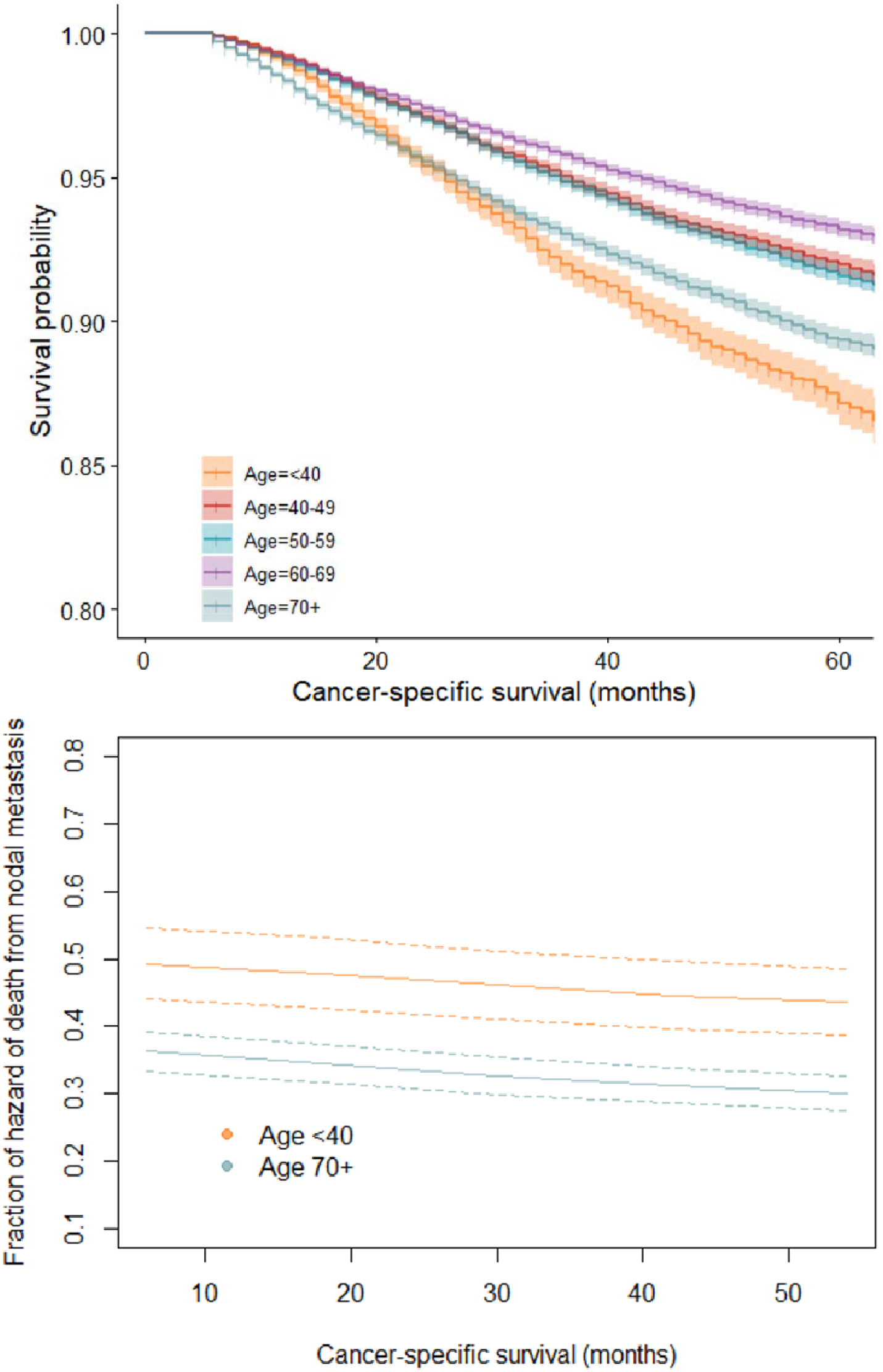
Univariate survival and NM-attributable fraction from adjusted models by selected age groups. A) Kaplan-Meier plot of survival by age group, SEER 2010-2017; B) Fraction of hazard of death due to NM for women under 40 years, and for women 70 years or more, SEER 2010-2017

## Discussion

In adult females younger than 40, there is considerable debate about whether breast cancer should be considered as a distinct disease. Women under 40 have consistently poor survival outcomes across various study populations [12,13,23–26] yet the reasons for this are not fully explained. The present study showed that the odds of NM at diagnosis have a gradient relationship to patient age of onset. This age-decreasing slope is most prominent for hormone receptor-positive subtypes. Findings from survival analysis confirmed that women under 40 have a hazard of death from breast cancer equal to or greater than the next-poorest group, women 70 years or older. Furthermore, the hazard of death attributable to NM was higher in women under 40 years, suggesting that NM has a stronger causal role in breast cancer mortality for younger women as compared to late age of diagnosis survival factors.

Previous studies have suggested that a higher incidence of the HR-/HER2+ subtype for young women with breast cancer partially explains their more aggressive disease [23–25]. Indeed, although our analysis showed a higher proportion of women under 40 years old having the HR-/HER2+ subtype, there was no significant difference in the odds of NM and age among the HR-/HER2+, and HR+ subtypes. However, a study, using a subset of the SEER data shows that, regardless of patients age, the HR+/HER2-breast cancer subtype has a higher rate of lymph node involvement at diagnosis than the triple-negative subtype [11]. From this study, NM at diagnosis does not appear to be related to HR-/HER2+ aggressiveness in young age of diagnosis. Furthermore, this observation may be of importance in considering whether prior research examining the link between the HR-/HER2+ subtype and nodal metastasis was adequately controlled for age at diagnosis.

The level of generalizability of the SEER 2010-2017 receptor-only subset of data may be questioned as a liability for analysis. We evaluated how representative subset data was using the complete, 42-year data. For all the variables common between datasets, both measures of association and measures of effect were consistent across data. Furthermore, we examined whether there was a time trend in proportion of NM positive patients at diagnosis in relation to age group. Interestingly, we found that, for women <40 years, the proportion of NM at diagnosis vs. non-NM remained stable across time at approximately 50%. In women aged 70+, there was a marked split of NM proportion from 50% starting in 1980, decreasing to ∼25% by 2008 (Figure 3).

**Figure 3.**
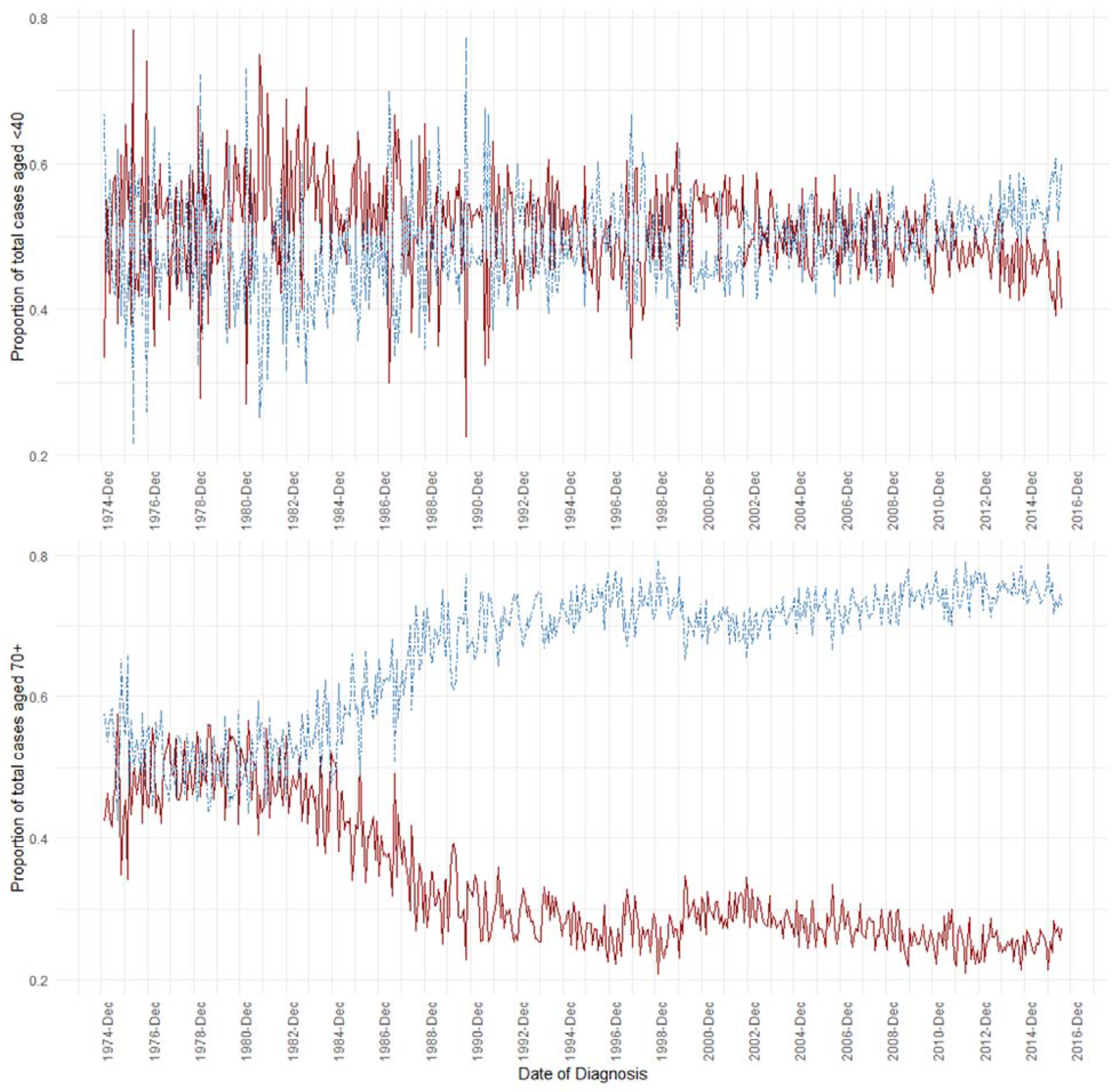
Lymph node metastasis (NM) as a proportion of total cases by month from 1975-2017, for ages under 40 years (top) and 70 years or older (bottom). NM in red, non-NM in blue.

The concordance of IHC-based receptor subtype with intrinsic molecular subtype has been shown to vary by subtype. Luminal B (HR+/HER2+) is the worst performing subtype in measures of both concordance and accuracy across several studies [27,28], suggesting that the relationship between young age of onset, NM, and interplay between estrogen/progesterone hormone receptors and HER2 should be studied further with a thorough consideration of molecular subtype classification. In contrast, previous research has shown the concordance and accuracy between both luminal A (HR+/HER2-) and triple-negative (HR-/HER2-) to be the highest among subtypes [29], suggesting that the results found in our study for these subtypes are likely not due to misclassification.

Lastly, there is also a question of consistency in measurement of variables over time. Perhaps the phenomenon of aggressive disease for younger women with breast cancer is influenced by changes in screening and diagnostic practices. From the standpoint of the association between age of diagnosis and NM, we included year of diagnosis as a predictor variable for both sets used for analysis. There was no confounding effect of the year of diagnosis upon the association between NM and age of diagnosis.

Involvement of lymph nodes is a key component in decisions for postoperative therapy, particularly radiation therapy, because clinicians evaluate need for lymph node radiation to minimize toxicity of treatment. Therefore, our results showing that the higher incidence of NM in young (<40 years) HR+ breast cancer groups (HR+/HER2- and HR+/HER2+) is clinically relevant. This relationship between NM and age of diagnosis is clear in women with luminal tumors, and may explain observations from previous studies linking the luminal subtype, young age of diagnosis, and poor prognosis [8]. Thus, our findings may aid in identifying aggressive disease in young women with luminal disease.

## Conclusions

Our results suggest that a predisposition towards more severe breast cancer for women with younger age of diagnosis is driven in part by NM. This relationship between NM and age of diagnosis for women with luminal tumors is strong and may be related to a poor prognosis. These findings also have implications in identifying high-risk, young HR+ groups (HR+/HER2- and HR+/HER2+) of breast cancer patients for aggressive therapy.

## Data Availability

All data is publicly available in SEER*stat database

https://seer.cancer.gov/seerstat/download/

## List of abbreviations

NM: nodal metastasis
SEER: Surveillance, Epidemiology, and End Results
TNM: tumor, node, and metastasis
HR: hormone receptor
ER: estrogen receptor
PR: progesterone receptor
ERBB2/HER2: human epidermal growth factor receptor 2
TNBC: triple-negative breast cancer

## DECLARATIONS

### Ethics approval and consent to participate

This study used publicly available data, thus these factors are not applicable.

### Consent for publication

Not applicable

### Availability of data and materials

The datasets used and/or analyzed during the current study are available from the corresponding author on reasonable request.

### Authors’ contributions

UM and MB conceptualized the study; MB, SA, PB, AE, and H-GK collected the data. MB analyzed the data, and MB, SW, SS, and UM interpreted the data regarding breast cancer. MB was a major contributor for this study; all authors contributed to writing the manuscript. All authors read and approved the final manuscript.

## Acknowledgements

This study was supported, in part, by grant 5U54CA118948 (NCI/NIH) and institutional/departmental impact funds to UM, and by CA047888 (NCI/NIH), the UAB Cancer Prevention and Control Training Program to MB (T32CA047888). We thank Dr. Donald Hill for his help in editing the manuscript.

## Authors’ information

MB, MPH, PhD-Postdoctoral Fellow of the Cancer Control and Prevention Program; PB, PhD-Researcher V; AE, MS-Graduate Student; H-GK, PhD - Researcher V; SW, MD, PhD-breast diagnostic pathologist, Professor of Pathology, and member of the O’Neal Comprehensive Cancer Center; SS, PhD, cancer epidemiologist, Professor of Epidemiology; and UM, MS, PhD, cancer translational researcher, Professor of Pathology, and member of the O’Neal Comprehensive Cancer Center.

